# Liver transplantation without graft ischemia in humans

**DOI:** 10.1101/2020.04.20.20065979

**Authors:** Zhiyong Guo, Qiang Zhao, Shanzhou Huang, Changjun Huang, Jian Zhang, Dongping Wang, Lu Yang, Maogen Chen, Linwei Wu, Zhiheng Zhang, Zebin Zhu, Linhe Wang, Caihui Zhu, Yixi Zhang, Yunhua Tang, Chengjun Sun, Wei Xiong, Yuekun Shen, Xiaoxiang Chen, Jinghong Xu, Tielong Wang, Yi Ma, Anbin Hu, Yinghua Chen, Xiaofeng Zhu, Jian Rong, Changjie Cai, Fengqiu Gong, Xiangdong Guan, Wenqi Huang, Dicken Shiu-Chung Ko, Xianchang Li, Jiefu Huang, Weiqiang Ju, Xiaoshun He

## Abstract

**BACKGROUND:** Ischemia-reperfusion injury is considered an inevitable event that compromises posttransplant outcomes. Numerous treatments have been proposed to reduce its impact. However, most of them have had limited success, as none of them can completely avoid graft ischemia.

**METHODS:** Ischemia-free liver transplantation (IFLT) comprises surgical techniques to enable continuous oxygenated blood supply to brain-dead donor livers during procurement, preservation and implantation using normothermic machine perfusion technology. In this nonrandomized study, 38 donor livers were transplanted using IFLT and were compared to 130 livers procured and transplanted using a conventional procedure (CLT).

**RESULTS:** One patient (2.6%) suffered early allograft dysfunction in the IFLT group, compared with 43.8% of patients in the CLT group (absolute risk difference, 41.2 percentage points; 95% confidence interval, −31.3, −51.1). The median (range) peak aspartate aminotransferase levels within the first week (336, 149-4112 vs. 1445, 149-25083 U/L, P<0.001), and the median (range) total bilirubin levels on day 7 (2.11, 0.68-12.47 vs. 5.11, 0.56-51.97 mg/dL, P<0.001) posttransplantation were much lower in the IFLT than in the CLT group. The IFLT recipients had less need for renal replacement therapy (2.6% vs. 16.9%, P=0.02), shorter median (range) intensive care unit stay (34, 12-235 vs. 43.5, 7-936 hours, P=0.003), and higher one-year recipient survival (97.4% vs. 84.6%, P=0.02) and graft survival (94.7% vs. 83.8%, P=0.04) rates than the CLT recipients. The extended criteria donor livers in IFLT yielded faster posttransplant recovery than the standard criteria donor livers in CLT.

**CONCLUSIONS:** IFLT provides a new approach to minimize ischemia-reperfusion injury and improve post-transplant outcomes.

**Clinical trial registry:** This trial is registered with http://www.chictr.org.cn, number ChiCTR-OPN-17012090.

## INTRODUCTION

Ischemia-reperfusion injury (IRI) has been considered an inevitable event in all conventional transplant procedures. IRI leads to a number of complications, such as early allograft dysfunction (EAD), primary nonfunction (PNF) and an increased incidence of allograft rejection in liver transplantation.^1^ Moreover, due to the severe organ shortage, organs from extended criteria donors (ECDs) are widely used worldwide.^2,3^ However, ECD organs are more vulnerable to IRI, leading to a higher risk of morbidity and mortality than is seen with standard criteria donor (SCD) organs.^4^ Therefore, IRI is a major obstacle to improved transplant outcomes and increased organ utilization.

Great efforts have been made to reduce IRI over the years, including ischemic preconditioning, the use of therapeutic gases, pharmacological interventions, stem cell therapy and gene therapy.^5^ However, limited success has been observed because none of these methods is able to reduce graft ischemia. As an alternative to standard static cold storage (SCS), *ex situ* normothermic machine perfusion (NMP) can provide a normothermic oxygenated blood supply to the organs. There are currently two modes of NMP, namely post-SCS (or “end-ischemic”) NMP and preservation NMP.^6^ Although both of them can reduce the cold ischemia time of grafts, they are not able to avoid procedures leading to graft ischemia during organ procurement, preparation and implantation.^7-12^

To completely avoid graft ischemia from retrieval in the donor to implantation in the recipient, we established a novel procedure called ischemia-free liver transplantation (IFLT) during which liver grafts were procured, preserved and implanted under continuous NMP.^13^ In the current study, we assessed the efficacy and safety of IFLT versus conventional liver transplantation (CLT) in patients with end-stage liver disease.

## METHODS

### PATIENTS AND STUDY DESIGN

In this single-center, nonrandomized trial, all donation after brain death (DBD) donors, aged 12-60 years, were eligible for inclusion. As the NMP device used in this study was not transportable, only organs from donors located in our own hospital were included. All donors were from the voluntary citizen-based organ donation system, and the organs were allocated through the China Organ Transplant Response System (COTRS).

All adult recipients (>18 years) on the waiting list of our hospital for a primary whole liver transplant with end-stage liver disease were potentially eligible, except those undergoing combined organ transplantation, multivisceral transplantation, split liver transplantation, and ABO-blood group incompatible transplantation. Eligible patients were approached for consent to receive IFLT, when the NMP device and perfusionists were available. If informed consent was provided, the patients underwent IFLT. Liver transplants using a conventional procedure at our center during the same period (from January 1st, 2017 to March 12th, 2019) with the same donor and recipient inclusion criteria, were used as contemporaneous comparators (CLT group).

After transplantation, standardized posttransplant care was provided in both groups, including fluid management, antibiotic and anti-hepatitis B virus (HBV) prophylaxis, immunosuppression, and surveillance ultrasonography. All the patients were followed up for one year posttransplantation. The study protocol (ChiCTR-OPN-17012090) was approved by the Ethical Committee of The First Affiliated Hospital, Sun Yatsen University.

## TRANSPLANT PROCEDURE

### IFLT procedure

Fig.1 and video 1 show the technical details of IFLT with multiple donor organ procurement.

**Figure 1.**
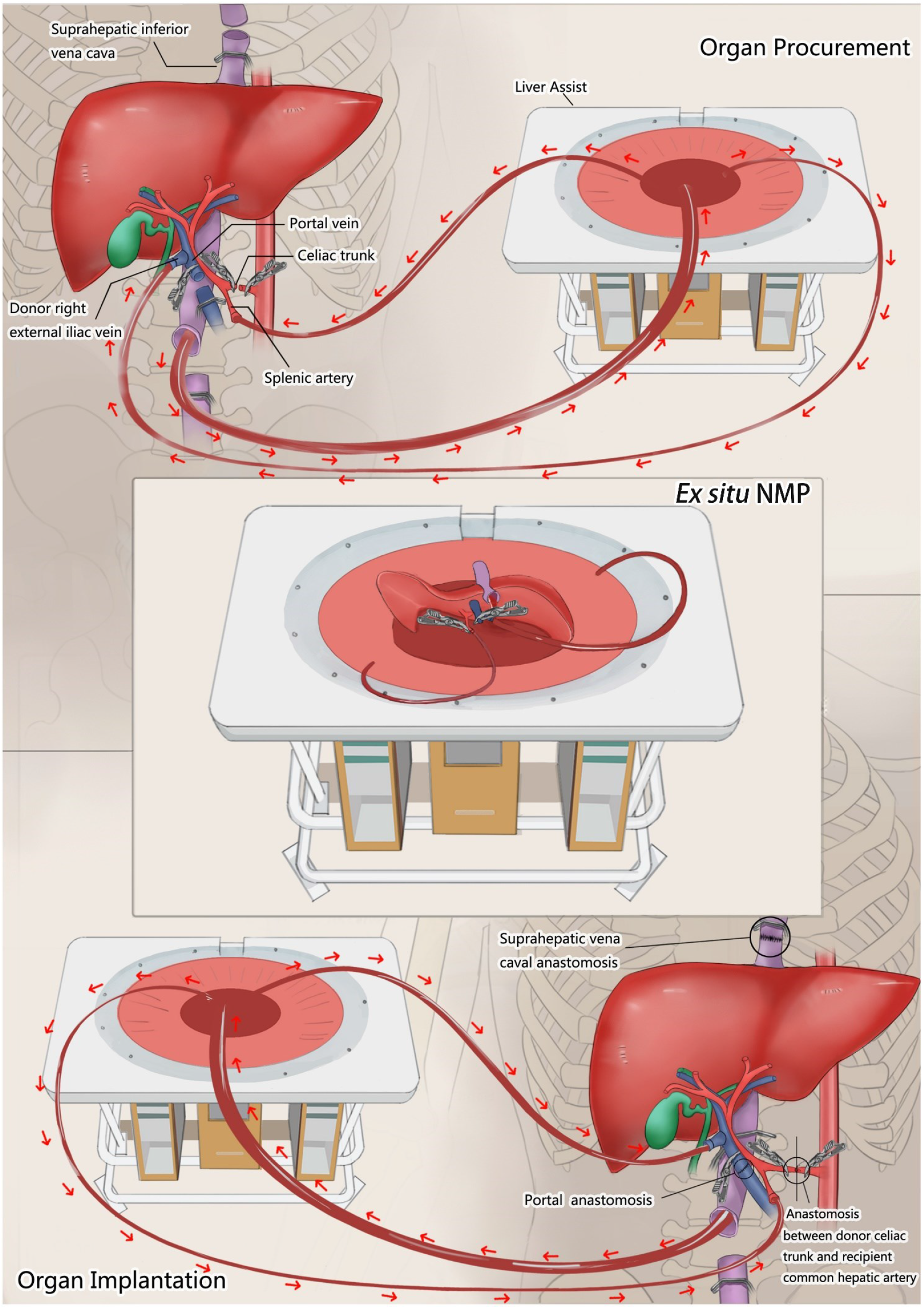
The Ischemia-free Liver Transplant Procedure. Liver procurement, *ex situ* preservation and implantation under normothermic machine perfusion (NMP) using the Liver Assist device with cannulation of the donor infrahepatic vena cava, interposition vein (right external iliac vein) on the portal vein, and splenic artery.

#### Organ procurement

After the liver was fully mobilized, a 12 Fr cannula was inserted into the splenic artery or gastroduodenal artery without interruption of the arterial supply to the liver from the celiac artery. A 32 Fr cannula was placed in the infrahepatic inferior vena cava for outflow and connected to the organ reservoir of the NMP device (Liver Assist, Organ Assist, Groningen, The Netherlands). A 24 Fr cannula, which was connected to the portal vein line of the device, was inserted into the portal vein via an interposition vein (the donor right external iliac vein). The arterial cannula was then connected to the arterial line of the device. After the *in situ* NMP circuit was established and perfusion began, the liver was procured and moved to the organ reservoir of the Liver Assist.

#### Ex-situ machine preservation

Once on the Liver Assist device, the liver underwent *ex situ* NMP. Livers were considered suitable for transplantation if they met all the following criteria during *ex situ* NMP: (i) the livers produced bile, (ii) the lactate level decreased to < 2.0 mmol/L within 90 min, (iii) the perfusate pH value was greater than 7.30, (iv) the arterial flow was greater than 150 ml/min and the portal venous flow was greater than 500 ml/min, and (v) the graft had a homogeneous appearance with soft consistency of the parenchyma. The NMP settings and perfusate composition were showed in our previous report.^13^

#### Organ implantation

After the hepatectomy of the diseased liver was finished, the donor liver was moved from the reservoir to the recipient peritoneal cavity. Liver implantation was performed using a bicaval or piggy-back technique. As a result of continuous *in situ* NMP of the liver via the splenic artery and interposition vein on the portal vein, all vascular anastomoses were conducted without interruption of blood supply to the graft. After graft revascularization, NMP was stopped and the cannulas were removed.

### CLT procedure

After the standard *in situ* cold flushing procedure, the liver was retrieved and placed in ice-cold University of Wisconsin (UW) solution and stored on ice. Back-table preparation was conducted under a standard procedure prior to implantation. After removal of the diseased liver, the donor liver was transferred to the abdominal cavity. Following anastomosis of the inferior vena cava and portal vein, the vessels were reopened to restore the blood supply of the allograft. Then, the hepatic artery and bile duct were anastomosed successively.

### GROUPING

Based on the donor types, the IFLT and CLT groups were divided into SCD and ECD subgroups. In this study, we characterized a graft as being from an ECD if at least one of the following criteria was met: (1) donor age > 60 years; (2) hypernatremia (serum Na^+^ > 165 mmol/L); (3) > 30% macrovesicular steatosis by biopsy; (4) donor serum aspartate aminotransferase (AST) or alanine aminotransferase (ALT) >1,000 IU/L or total bilirubin (Tbil) > 3 mg/dL at the time of organ offer; (5) or cold ischemia time (CIT) ≥ 12 hours.

### FOLLOW UP OF PATIENT OUTCOMES

The patients were followed up for one year posttransplantation. EAD was defined by the presence of one of peak AST or ALT levels >2000 U/L within the first 7 days, international normalized ratio (INR) >1.6 or Tbil >10 mg/dL on day 7 posttransplantation, with exclusion of anastomostic biliary stricture and hepatic artery thrombosis (HAT).^14^ Other outcomes included PNF, defined as graft failure immediately after transplantation requiring urgent retransplantation or leading to patient death;^15^ biliary complications including non-anastomotic biliary stricture (NAS) defined by Carlijn et al;^16^ vascular complications; clinical acute rejection; need for renal replacement therapy (RRT) within 30 days; length of stay in the intensive care unit (ICU); length of posttransplant stay in the hospital; one-year patient survival and graft survival.

### STATISTICAL ANALYSIS

The results of donor and recipient characteristics, as well as the results of clinical outcomes, are expressed in the median (range) or the mean ± standard deviation (SD) for continuous parameters and in percentages for nominal parameters. Continuous parameters were compared with a 2-tailed Student’s T-test or a 2-tailed Mann-Whitney nonparametric test, while Fisher’s exact test was used to compare categorical parameters. For categorical outcomes, absolute risk differences between the IFLT and CLT groups was calculated using exact unconditional methods based on the Farrington-Manning score statistic, expressed as percentage points with 95% confidence intervals (CIs). The linear graphs of perfusion parameters, blood gas analysis and bile examination are presented as the median and range. Subgroup analyses were performed for the donor types (ECD and SCD). P<0.05 was considered to be statistically significant. The data analysis was performed using Statistical Product and Service Solutions (SPSS) 22.0 (IBM, New York, USA).

## RESULTS

### DONOR AND RECIPIENT CHARACTERISTICS

During the study period, 412 donor livers were allocated to our center for transplantation. A total of 168 donor livers meeting both donor and recipient inclusion criteria were successfully transplanted. Thirty-eight patients underwent IFLT and 130 patients underwent CLT (Fig. 2). Table 1 summarizes the donor and recipient characteristics. There was no statistical difference in the donor sex, age, body mass index (BMI), causes of death and donor types between the two groups. There were 12 (31.6%) and 29 (22.3%) ECD organs in the IFLT and CLT groups, respectively (Table S1 in the Supplementary Appendix). The organ utilization rates of kidneys, lungs and pancreas were comparable in the two groups, while the utilization rate of hearts was higher in the IFLT group than in the CLT group (26.3% vs. 10.8%, P = 0.002) (Table S2 in the Supplementary Appendix). The renal transplant outcomes were comparable between the two groups (Table S3 and Fig. S1 in the Supplementary Appendix). There was no significant difference in the recipient age, sex, model for end-stage liver disease (MELD) score, or primary diagnosis of liver diseases between the two groups.

**Table 1.**
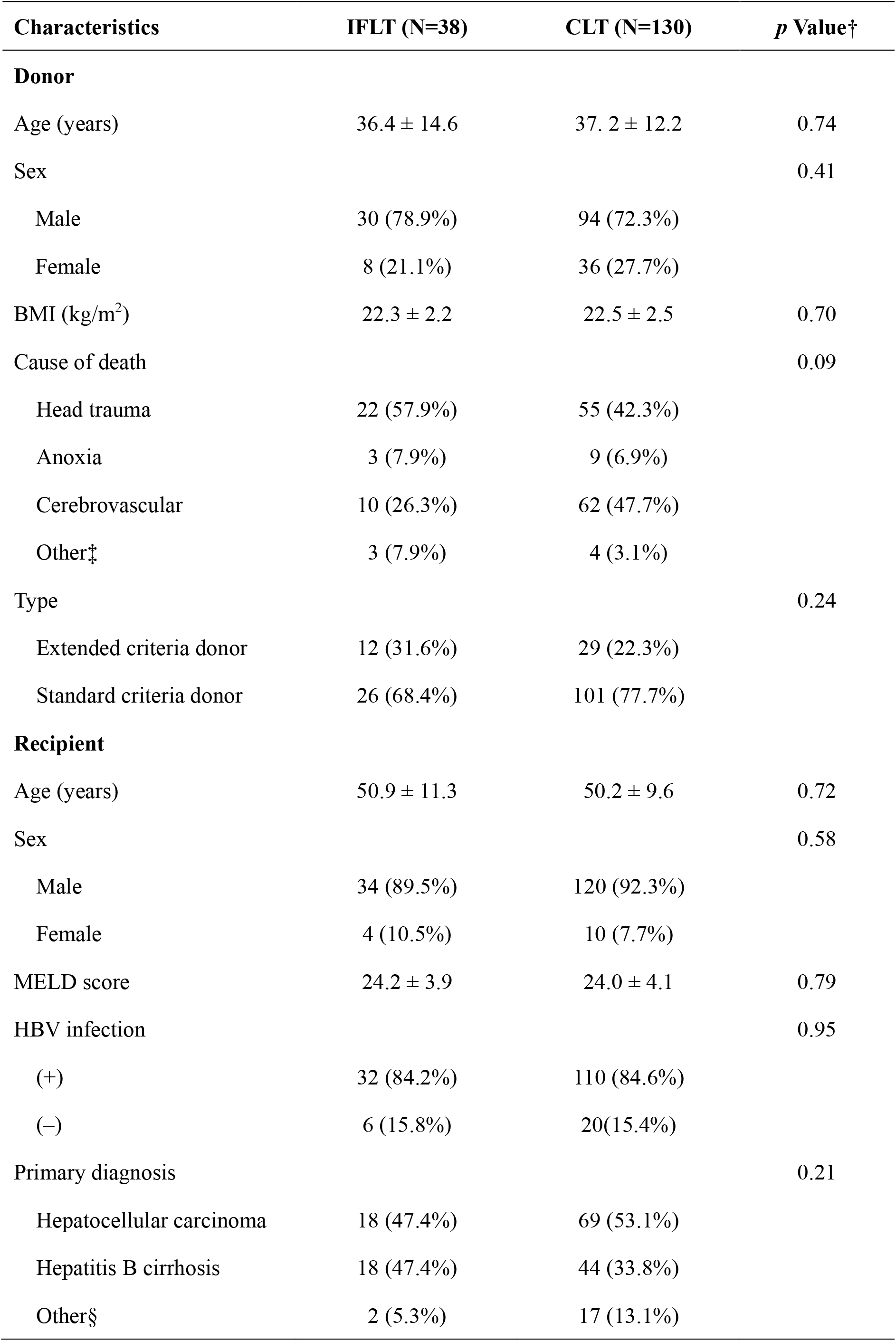
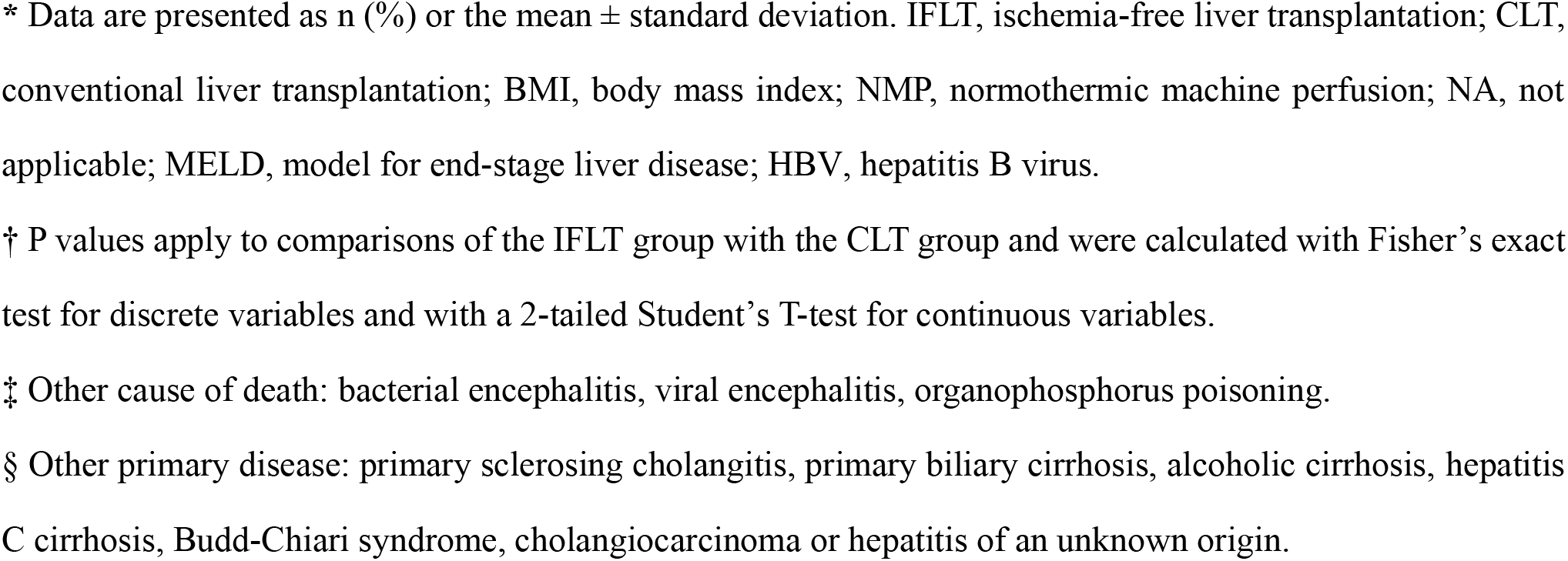
Baseline demographic and clinical characteristics of the donors and recipients.*

| Characteristics | IFLT (N=38) | CLT (N=130) | *p* Value† |
| --- | --- | --- | --- |
| **Donor** |  |  |  |
| Age (years) | 36.4 ± 14.6 | 37. 2 ± 12.2 | 0.74 |
| Sex |  |  | 0.41 |
| Male | 30 (78.9%) | 94 (72.3%) |  |
| Female | 8 (21.1%) | 36 (27.7%) |  |
| BMI (kg/m<sup>2</sup>) | 22.3 ± 2.2 | 22.5 ± 2.5 | 0.70 |
| Cause of death |  |  | 0.09 |
| Head trauma | 22 (57.9%) | 55 (42.3%) |  |
| Anoxia | 3 (7.9%) | 9 (6.9%) |  |
| Cerebrovascular | 10 (26.3%) | 62 (47.7%) |  |
| Other‡ | 3 (7.9%) | 4 (3.1%) |  |
| Type |  |  | 0.24 |
| Extended criteria donor | 12 (31.6%) | 29 (22.3%) |  |
| Standard criteria donor | 26 (68.4%) | 101 (77.7%) |  |
| **Recipient** |  |  |  |
| Age (years) | 50.9 ± 11.3 | 50.2 ± 9.6 | 0.72 |
| Sex |  |  | 0.58 |
| Male | 34 (89.5%) | 120 (92.3%) |  |
| Female | 4 (10.5%) | 10 (7.7%) |  |
| MELD score | 24.2 ± 3.9 | 24.0 ± 4.1 | 0.79 |
| HBV infection |  |  | 0.95 |
| (+) | 32 (84.2%) | 110 (84.6%) |  |
| (−) | 6 (15.8%) | 20(15.4%) |  |
| Primary diagnosis |  |  | 0.21 |
| Hepatocellular carcinoma | 18 (47.4%) | 69 (53.1%) |  |
| Hepatitis B cirrhosis | 18 (47.4%) | 44 (33.8%) |  |
| Other§ | 2 (5.3%) | 17 (13.1%) |  |

\* Data are presented as n (%) or the mean ± standard deviation. IFLT, ischemia-free liver transplantation; CLT, conventional liver transplantation; BMI, body mass index; NMP, normothermic machine perfusion; NA, not applicable; MELD, model for end-stage liver disease; HBV, hepatitis B virus.
† P values apply to comparisons of the IFLT group with the CLT group and were calculated with Fisher's exact test for discrete variables and with a 2-tailed Student's T-test for continuous variables.
‡ Other cause of death: bacterial encephalitis, viral encephalitis, organophosphorus poisoning.
§ Other primary disease: primary sclerosing cholangitis, primary biliary cirrhosis, alcoholic cirrhosis, hepatitis C cirrhosis, Budd-Chiari syndrome, cholangiocarcinoma or hepatitis of an unknown origin.

**Figure 2.**
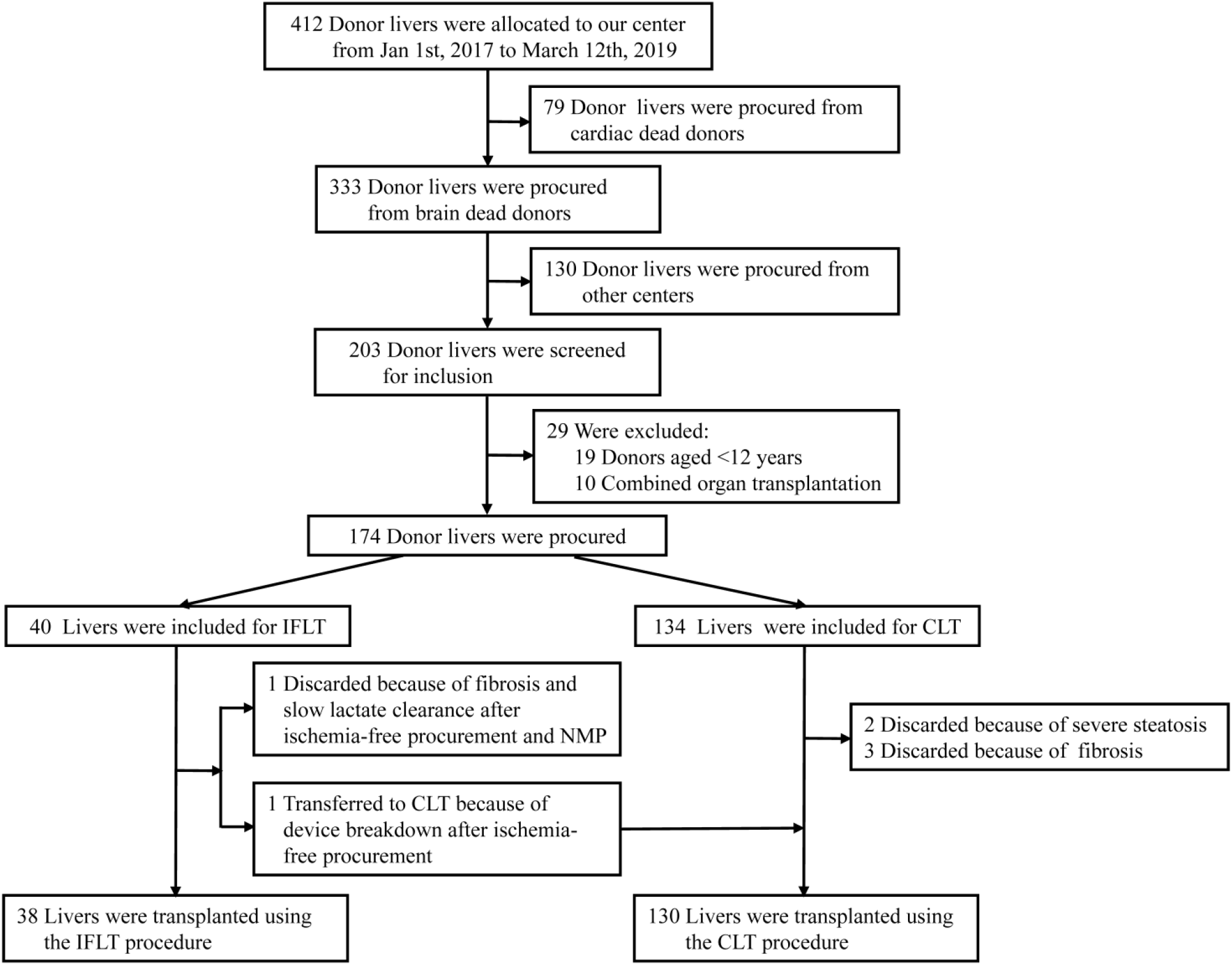
Screening, Selection, and Follow-Up of the Patients. A total of 412 donor livers that were screened from January 1st, 2017, to March 12th, 2019, 79 donation after cardiac death (DCD) livers and 130 livers from other centers were excluded, and 203 donation after brain death (DBD) livers were left. Nineteen livers from pediatric donors and 10 livers used for combined organ transplantation were excluded. Of the remaining 174 livers, 40 livers were included for ischemia-free liver transplantation (IFLT), and 134 livers were included for conventional liver transplantation (CLT). Of the 40 livers, one was discarded because of fibrosis and slow lactate clearance after ischemia-free procurement and NMP, and one was transferred to CLT because of device breakdown after ischemia-free procurement. Of the 134 livers for CLT, two were discarded because of severe steatosis and three were discarded because of fibrosis. Eventually, 38 livers were transplanted using the IFLT procedure and 130 livers were transplanted using the CLT procedure.

The anhepatic phase of the recipient operations were comparable between the two groups (P=0.97), although the median (range) duration of the recipient operations was shorter in the IFLT than in the CLT group (385, 238-515 vs. 445, 285-945 min, P<0.001). During transplantation, the mean (± SD) body temperature of the recipients was higher during an-hepatic phase (35.69 ± 0.70 vs. 34.66 ± 3.14°C, P= 0.006) and within one hour (36.30 ± 0.72 vs. 35.33 ± 0.96 °C, P<0.001) after graft revascularization in the IFLT than in the CLT (Fig. S2 in the Supplementary Appendix). There was no significant difference in intraoperative blood loss, use of red blood cells and fresh frozen plasma, as well as post-transplant INR, prothrombin time (PT) and fibrinogen (Fbg) levels between the two groups (Table S4 and Fig. S3 in the Supplementary Appendix).

### NMP PARAMETERS AND BIOCHEMICAL ANALYSIS OF THE PERFUSATE IN IFLT

A total of 40 donor livers were subjected to NMP. One liver was cooled down with 0-4°C UW solution and transferred to CLT because the hepatic artery pump of the device stopped running after the donor liver was moved from the donor peritoneal cavity to the organ reservoir. One liver was discarded because of slow lactate clearance and obvious fibrosis secondary to hepatitis B virus infection. Fig. S4 in the Supplementary Appendix shows the NMP parameters and biochemical analysis of perfusate of the 38 IFLT cases. The pressure and flow of both the portal vein and hepatic artery were stable throughout the entire IFLT procedure. The partial pressure of oxygen (pO_2_) were among 150-250 mmHg and partial pressure of carbon dioxide (pCO_2_) were among 35-45 mmHg. Biochemical analysis of the perfusate showed that the pH values were within normal ranges (7.35-7.45). The lactate level declined rapidly from the original level to less than 2.0 mmol/L. All the perfused livers continued producing bile during the whole procedure.

### REDUCED LIVER GRAFT INJURIES AFTER IFLT

The median (range) peak AST level within 7 days posttransplantation was lower in the IFLT group (336, 149-4,112 U/L) than in the CLT group (1,445, 149-25,083 U/L) (P<0.001). Similarly, the peak ALT level was much lower in the IFLT group (155, 48-3,474 U/L) than in the CLT group (693, 86-10,500 U/L) (P<0.001). In addition, the Tbil level on day 7 posttransplantation was lower in the IFLT group (2.11, 0.68-12.47 mg/dL) than in the CLT group (5.11, 0.56-51.97 mg/dL) (P<0.001) (Table 2). The IFLT group demonstrated lower cumulative levels of the liver injury markers of AST, ALT, Tbil and lactate dehydrogenase (LDH) in the early phase posttransplantation than the CLT group. The IFLT recipients had significantly lower gamma-glutamyl transpeptidase (GGT) and alkaline phosphatase (ALP) levels at one year posttransplantation than the CLT recipients (Fig. S5 in the Supplementary Appendix).

**Table 2.**
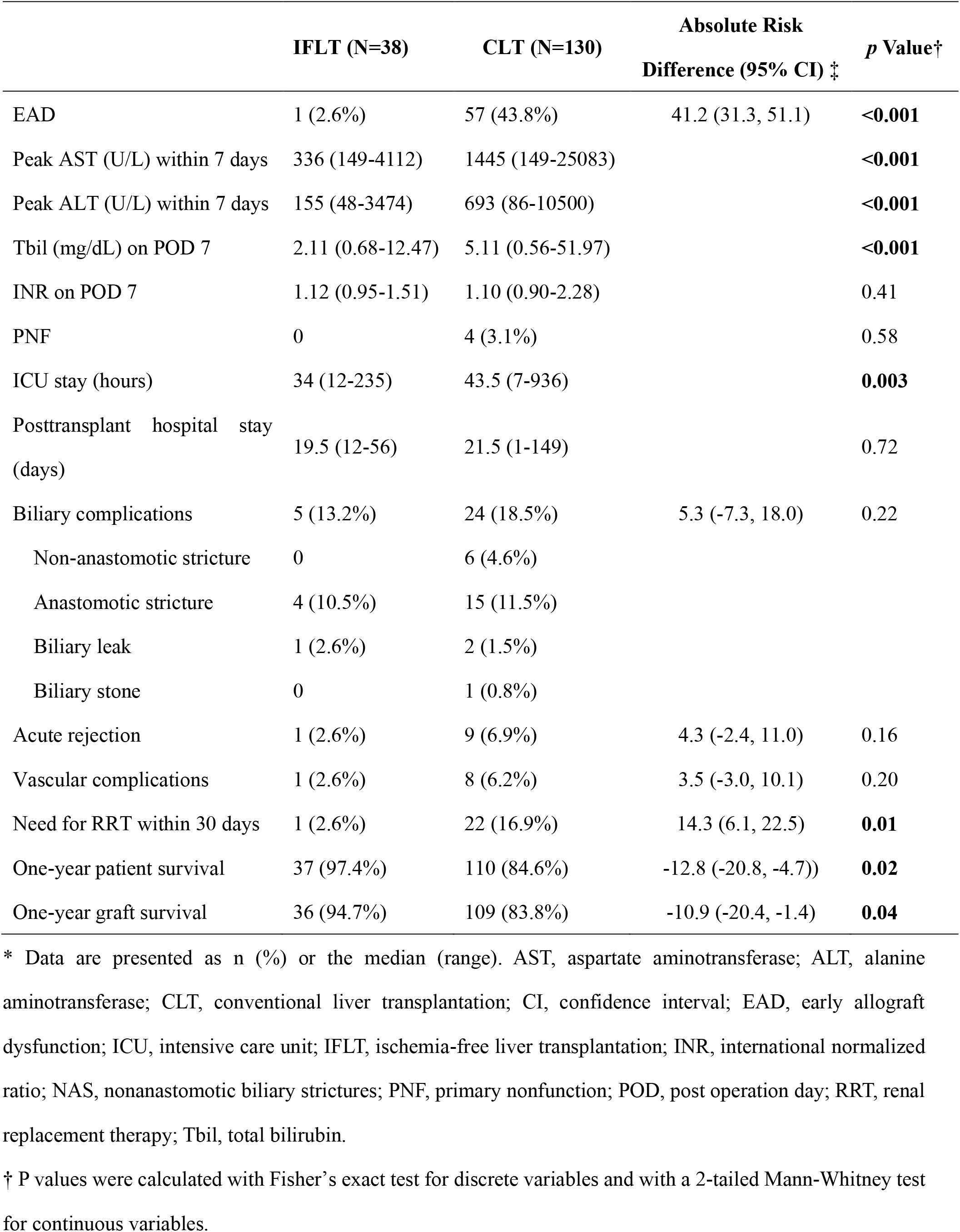
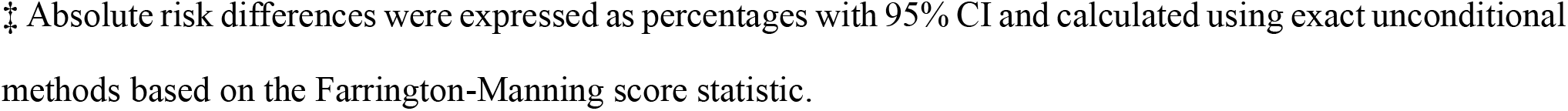
Outcomes in the IFLT and CLT groups.*

|  | IFLT (N=38) | CLT (N=130) | Absolute Risk<br>Difference (95% CI) ‡ | p Value† |
| --- | --- | --- | --- | --- |
| EAD | 1 (2.6%) | 57 (43.8%) | 41.2 (31.3, 51.1) | <0.001 |
| Peak AST (U/L) within 7 days | 336 (149-4112) | 1445 (149-25083) |  | <0.001 |
| Peak ALT (U/L) within 7 days | 155 (48-3474) | 693 (86-10500) |  | <0.001 |
| Tbil (mg/dL) on POD 7 | 2.11 (0.68-12.47) | 5.11 (0.56-51.97) |  | <0.001 |
| INR on POD 7 | 1.12 (0.95-1.51) | 1.10 (0.90-2.28) |  | 0.41 |
| PNF | 0 | 4 (3.1%) |  | 0.58 |
| ICU stay (hours) | 34 (12-235) | 43.5 (7-936) |  | **0.003** |
| Posttransplant hospital stay<br>(days) | 19.5 (12-56) | 21.5 (1-149) |  | 0.72 |
| Biliary complications | 5 (13.2%) | 24 (18.5%) | 5.3 (-7.3, 18.0) | 0.22 |
| Non-anastomotic stricture | 0 | 6 (4.6%) |  |  |
| Anastomotic stricture | 4 (10.5%) | 15 (11.5%) |  |  |
| Biliary leak | 1 (2.6%) | 2 (1.5%) |  |  |
| Biliary stone | 0 | 1 (0.8%) |  |  |
| Acute rejection | 1 (2.6%) | 9 (6.9%) | 4.3 (-2.4, 11.0) | 0.16 |
| Vascular complications | 1 (2.6%) | 8 (6.2%) | 3.5 (-3.0, 10.1) | 0.20 |
| Need for RRT within 30 days | 1 (2.6%) | 22 (16.9%) | 14.3 (6.1, 22.5) | **0.01** |
| One-year patient survival | 37 (97.4%) | 110 (84.6%) | -12.8 (-20.8, -4.7)) | **0.02** |
| One-year graft survival | 36 (94.7%) | 109 (83.8%) | -10.9 (-20.4, -1.4) | **0.04** |

\* Data are presented as n (%) or the median (range). AST, aspartate aminotransferase; ALT, alanine aminotransferase; CLT, conventional liver transplantation; CI, confidence interval; EAD, early allograft dysfunction; ICU, intensive care unit; IFLT, ischemia-free liver transplantation; INR, international normalized ratio; NAS, nonanastomotic biliary strictures; PNF, primary nonfunction; POD, post operation day; RRT, renal replacement therapy; Tbil, total bilirubin.
† P values were calculated with Fisher's exact test for discrete variables and with a 2-tailed Mann-Whitney test for continuous variables.
‡ Absolute risk differences were expressed as percentages with 95% CI and calculated using exact unconditional methods based on the Farrington-Manning score statistic.

### IMPROVED POSTTRANSPLANT OUTCOMES AFTER IFLT VERSUS CLT

One (2.6%) patient developed EAD in the IFLT group, while 57 (43.8%) patients in the CLT group suffered EAD (P<0.001) (Table 2). The majority (32/58, 55.2%) of the EAD cases met the Tbil criteria (Table S5 in the Supplementary Appendix). No patient suffered PNF in the IFLT group, while four cases of PNF occurred in the CLT group. The IFLT recipients had a shorter median (range) post-transplant ICU stay than the CLT recipients (34, 12-235 hours vs. 43.5, 7-936 hours, P<0.001). Twenty-two out of 130 (16.9%) recipients needed renal replacement treatment in the CLT group, compared with only one patient (2.6%) in the IFLT group (P=0.01). The incidence of biliary complications was 18.5% (24/130) with 6 NAS in the CLT group and 13.2% (5/38) with no NAS in the IFLT group (P=0.22). The incidences of vascular complications and rejection were comparable between the two groups. The patients in the IFLT group had a superior one-year patient survival when compared to those in the CLT group (97.4% versus 84.6%, P=0.02). Four patients died of PNF in the CLT group (Table S6 in the Supplementary Appendix). Moreover, the one-year graft survival was higher in the IFLT than in the CLT group (94.7% versus 83.8%, P=0.04).

### FASTER RECOVERY OF ECD LIVERS POSTTRANSPLANTATION FACILITATED BY IFLT

To further clarify the potential advantages of IFLT in using ECD livers, we performed a subgroup analysis by comparing the liver function test results of the IFLT_ECD subgroup with the IFLT_SCD, CLT_SCD and CLT_ECD subgroups. The IFLT_ECD subgroup had a comparable post-transplant peak AST/ALT values and Tbil on POD 7 to the IFLT_SCD subgroup. Livers had lower median (range) peak AST (365, 149-4,112 vs. 1,325, 149-20,583 U/L, P<0.001) and ALT (259, 66-404 vs. 658, 86-10,500 U/L, P<0.001) values and Tbil values on POD 7 (1.65, 0.77-12.47 vs. 3.61, 0.56-37.08 mg/dL, P=0.04) levels in the IFLT_ECD than in the CLT_SCD subgroup. There was no statistical significance in INR on post-transplant day 7 between the four subgroups. The IFLT_ECD group had no incidence of EAD, which was significantly lower than than in both the CLT_SCD (40.6%, P=0.004) and CLT_ECD subgroups (55.2%, P=0.001) (Table S7 and Fig. S6 in the Supplementary Appendix).

## DISCUSSION

In all conventional transplant procedures, the oxygenated blood supply to the organ is completely suspended during procurement, SCS preservation and implantation, which leads to ischemic damage to the donor organs. The restoration of oxygenated blood supply (graft revascularization) after ischemia exacerbates the initial cellular damage; this process is known as IRI.^1^ Based on the assumption that IRI is an inevitable event in organ transplantation, all proposed methods have attempted to reduce IRI instead of to avoid it. In contrast to existing methods, the IFLT procedure was developed to completely avoid graft ischemia. The results of this first clinical trial showed that IFLT can reduce posttransplant complications and improve one-year patient and graft survival compared with CLT.

In China, the deceased organ donation system was formally established in 2015.^17^ The donors often suffer hypotension, hypoxia and <u>anemia.</u> The incidence of EAD is 36.4-54.8% according to the national database, which is much higher than that reported by Western countries.^7,18^ Consistent with this, 43.8% of recipients suffered EAD in the CLT group. In contrast, only one patient developed EAD in the IFLT group, which was thought to be due to a massive liver hematoma after biopsy. Notably, four patients died of PNF (3.1%) in the CLT group, while no patient suffered PNF in the IFLT group. In addition, no NAS occurred in the IFLT group, while six patients developed NAS in the CLT group. Furthermore, the Tbil, GGT and ALP levels were much lower one year posttransplantation in the IFLT than in the CLT group, suggesting the protective effects of IFLT on the bile ducts. Collectively, these results show that IFLT is able to prevent the occurrence of major IRI-related complications.

Recently, the definition of EAD has been challenged as an endpoint in studies related to NMP because the liver enzyme might be “washed out” during perfusion.^19^ In the current study, the AST/ALT concentrations in the perfusate were stably low. In addition, more than half of EAD cases were defined by the Tbil criteria in this study. These results suggest that the “washed out” effect cannot explain the difference in the incidence of EAD between the two groups. In addition, it has been reported that EAD cannot predict graft survival.^20^ However, in the current study, patients with EAD had much lower patient and graft survival rates than those without EAD in the CLT group (Fig. S7 in the Supplementary Appendix). Moreover, the incidence of peak AST > 5,000 U/L, which is a predictor of inferior graft survival,^21^ was significantly lower in the IFLT than in the CLT group (0 vs. 13.1%, P<0.001). Therefore, it is of clinical relevance to reduce EAD, at least based on the current study.

To reduce the IRI of ECD organs, various types of machine perfusion have been used in clinical practice, such as hypothermic machine perfusion (HMP), hypothermic oxygenated perfusion (HOPE), NMP, subnormothermic machine perfusion (SNP) and controlled oxygenated rewarming (COR).^6,7,22-27^ These novel preservation methods are potentially able to assess graft viability and improve transplant outcomes. Nevertheless, the organ procurement and implantation techniques are the same as CLT, even when these techniques are used. The grafts still suffer ischemia and subsequent IRI. By taking advantage of the tri-branch structure of the portal vein, celiac artery and posthepatic inferior vena cava, IFLT enables a continuous warm blood supply for the donor livers by switching between *in vivo* blood perfusion and NMP during organ procurement and implantation, which has the potential to avoid IRI. The first case of IFLT successfully transplanted a donor liver with 85-90% macrovesicular hepatosteatosis,^13^ which is far beyond the accepted macrovesicular hepatosteatosis grade (<60%) in the majority of transplant centers worldwide.^28^ In the current study, by using IFLT, the ECD livers yielded comparable graft function recovery to the SCD livers and a much faster recovery than the SCD livers in the CLT group. Therefore, IFLT is a promising approach to increase organ utilization.

Our group has developed a multiple organ procurement protocol when IFLT is conducted. There was no significant difference in the organ recovery rate or posttransplant outcomes of the nonliver grafts in the current study. Importantly, it has been shown that NMP is safe and feasible in lung, heart and kidney preservation. Therefore, it is possible that the concept of ischemia-free organ transplantation can also be adopted in these organs. The techniques of ischemia-free kidney transplantation have been reported by our center early this year.^29^ Notably, one liver intended to undergo IFLT was transferred to SCS due to a defect in NMP disposable. Another group also reported technical issues with NMP.^30^ Therefore, a cold UW solution should be prepared in all cases of IFLT in case of problems, and a well-trained perfusionist is required for the safety of the technique. There are limitations in the current study. First, a nontransportable machine perfusion device was used in this study, and the livers were all locally procured in both groups. A simplified technique with a portable device is under development in our center to enable distant procurement. Second, this was the first clinical trial to assess the safety and efficacy of IFLT in humans. The nonrandomized design of this study might lead to patient selection bias, although no significant difference in either donor or recipient characteristics was found between the two groups. A randomized controlled trial (ChiCTR1900021158) is ongoing in our center to confirm the above findings. Finally, although IFLT represents an extreme example of NMP, the potential benefits of IFLT over liver transplantation using preservation NMP, particularly in ECD livers, are yet to be defined.

In conclusion, IFLT can provide an efficient approach to minimize IRI and improve transplant outcomes, which might change current practice in liver transplantation. Further development and modification of the technique might improve its safety and efficacy. In addition, randomized controlled studies comparing the distinct types of machine perfusion techniques should be conducted to define their respective advantages and disadvantages in different clinical conditions.

## Data Availability

All data needed to evaluate the conclusions in the paper are present in the paper or the supplementary materials.

## ACKNOWLEDGEMENTS

Supported by grants as follows: the National Natural Science Foundation of China (81970564, 81471583 and 81570587), the Key Clinical Specialty Construction Project of National Health and Family Planning Commission of the People’s Republic of China, the Guangdong Provincial Key Laboratory Construction Projection on Organ Donation and Transplant Immunology (2013A061401007, 2017B030314018), Guangdong Provincial international Cooperation Base of Science and Technology (Organ Transplantation) (2015B050501002), Guangdong Provincial Natural Science Funds for Major Basic Science Culture Project (2015A030308010), Guangdong Provincial Natural Science Funds for Distinguished Young Scholars (2015A030306025), Special support program for training high level talents in Guangdong Province (2015TQ01R168), Pearl River Nova Program of Guangzhou (201506010014), Science and Technology Program of Guangzhou (201704020150), Sun Yat-sen University Young Teacher Key Cultivate Project (17ykzd29) and “Elite program” specially supported by China organ transplantation development foundation.

We thank all members of the organ procurement organization, surgical intensive care unit, transplant anesthesia team and cardiopulmonary bypass team.

